# The origin and early spread of SARS-CoV-2 in Europe

**DOI:** 10.1101/2020.06.10.20127738

**Authors:** Sarah A. Nadeau, Timothy G. Vaughan, Jérémie Sciré, Jana S. Huisman, Tanja Stadler

## Abstract

The investigation of migratory patterns of the SARS-CoV-2 pandemic before border closures in Europe is a crucial first step towards an in-depth evaluation of border closure policies. Here we analyze viral genome sequences using a phylodynamic model with geographic structure to estimate the origin and spread of SARS-CoV-2 in Europe prior to border closures. Based on SARS-CoV-2 genomes, we reconstruct a partial transmission tree of the early pandemic, including inferences of the geographic location of ancestral lineages and the number of migration events into and between European regions. We find that the predominant lineage spreading in Europe has a most recent common ancestor in Italy and was probably seeded by a transmission event in either Hubei or Germany. We do not find evidence for preferential migration paths from Hubei into different European regions or from each European region to the others. Sustained local transmission is first evident in Italy and then shortly thereafter in the other European regions considered. Before the first border closures in Europe, we estimate that the rate of occurrence of new cases from within-country transmission was within the bounds of the estimated rate of new cases from migration. In summary, our analysis offers a view on the early state of the epidemic in Europe and on migration patterns of the virus before border closures. This information will enable further study of the necessity and timeliness of border closures.

**Significance Statement:** We estimate the origin and spread of SARS-CoV-2 in Europe prior to border closures based on viral genome sequences using a phylodynamic model with geographic structure. We confirm that the predominant European outbreak most likely started in Italy and spread from there. This outbreak was probably seeded by a transmission event in either Hubei or Germany. In particular, we find that before the first border closures in Europe, the rate of new cases occurring from within-country transmission was within the estimated bounds on the rate of new migration cases.

## Introduction

In response to the pandemic potential of the SARS-CoV-2 virus, many nations closed their borders in order to curb the virus’ spread (1). These closures continue to incur high economic and social costs. To weigh the relative costs and benefits of border closures, it will be important to understand the efficacy of these policies. At the early stages of an outbreak, border closures can delay a pathogen’s arrival, thereby giving countries additional time to prepare (2). However, the success of this strategy depends on timely implementation and a good knowledge of where the pathogen is already circulating. To evaluate the efficacy of border closures in limiting the spread of SARS-CoV-2, it is important to reconstruct the timeline of the early international spread of the virus, before such policies were implemented.

In this analysis, we aim to estimate the early patterns of SARS-CoV-2 transmission into and across Europe. We also address the more specific question of where the predominant SARS-CoV-2 lineage circulating in Europe originated. We hope that by addressing these questions we can inform further analysis of the efficacy of border closures as a strategy to combat SARS-CoV-2.

The SARS-CoV-2 virus was identified as the cause of an epidemic in Wuhan, China in late 2019 (3). The epidemic in Wuhan was reported to the WHO on 31 Dec. 2019 and within one month, SARS-CoV-2 was confirmed to have spread to 19 additional countries (4). By the end of February 2020, the virus was detected in all WHO regions (5). Currently, several lineages of the SARS-CoV-2 virus are circulating across the globe. The intermixing of these lineages in different countries and regions suggests that the virus has been transmitted across borders many times (6).

Here we focus on estimating the early introductions of SARS-CoV-2 into Europe and the virus’ migration across European borders. Through national surveillance efforts, the first COVID-19 cases in Europe were detected in France on 24 Jan. 2020 and in Germany on 28 Jan. 2020 (7, 8). Of the 47 cases detected in Europe by 21 Feb. 2020, 14 were infected in China, 14 were linked to the initial cases in Germany, 7 were linked to the initial cases in France, and 12 were of unknown origin (7). In addition to the unknown sources of transmission, some early introductions may not have been detected. This is especially probable given that a significant proportion of infected individuals are likely to be asymptomatic (9). In summary, it is difficult to draw firm conclusions about the source, number, and timing of SARS-CoV-2 introductions into Europe based on confirmed case data alone.

Viral genomes are an important secondary source of information on outbreak dynamics. If viruses acquire mutations on the same timescale as an outbreak, these mutations can provide information about past transmission events. Phylodynamic methods couple a model of viral evolution describing the mutational process to an epidemiological model describing the transmission process. By fitting the combined model to viral genomes sampled from a cohort of infected individuals, we can infer the evolutionary and epidemiological model parameters. Here we fit a phylodynamic model with geographic structure to SARS-CoV-2 genomes from Hubei, China and several European countries before the first borders were closed in these regions. We co-infer the transmission tree linking these sequences, the geographic location of ancestral lineages, migration rates of infected individuals between regions, the effective reproductive number, and the proportion of no-longer infectious cases sequenced in each region.

In addition to these inferences, we specifically focus on estimating the geographic origin of the predominant SARS-CoV-2 lineage in Europe. This lineage is defined by a characteristic amino acid substitution at position 314 in the ORF1b gene from proline to leucine and was provisionally named the “A2a” lineage by the Nextstrain team. In the more dynamic, tree-based nomenclature suggested by (10), this lineage corresponds to the “B.1” lineage described as “a large lineage that roughly corresponds to the large outbreak in Italy, and has since seeded many different countries” (11). As of Apr. 1, 2020, two-thirds of the SARS-CoV-2 sequences collected in Europe belonged to this lineage and just 10% of sequences from the lineage were collected outside Europe (data from (12), lineages assigned using (13)). Here, we use the name A2a to refer to the group of SARS-CoV-2 viruses defined by the ORF1b:P314L mutation.

Where the A2a lineage originated remains unclear. Its characteristic ORF1b mutation was found in some of the earliest confirmed COVID-19 cases in Italy, Switzerland, Germany, Finland, Mexico, and Brazil in late February (14, 15). Intriguingly, a late-January sample from a German case linked to business travel from Singapore shares a mutation in the S gene with the A2a lineage, but does not have the lineage-defining ORF1b mutation. This German sample is part of a smaller “A2” clade that is basal to the larger clade of A2a sequences (6). As a result, it was hypothesized that a German transmission cluster may have seeded the larger European outbreak (14–16). However, it was quickly pointed out that incomplete and biased sampling must be taken into account before this hypothesis can be rigorously addressed (14, 17, 18).

Phylodynamic models with geographic structure aim to account for such biases. Firstly, parameter estimates are generated by integrating over a distribution of potential phylogenies, which acknowledges that we cannot reconstruct the true transmission tree with certainty. Secondly, sampling parameters are allowed to differ between regions, which acknowledges that testing and sequencing resources vary across regions. Here, we fit a phylodynamic model with geographic structure to full-length SARS-CoV-2 genomes to (i) estimate the early patterns of SARS-CoV-2 spread into and across Europe, (ii) weigh genomic evidence for competing hypotheses about the geographic origin of the predominant A2a lineage in Europe, (iii) report on the epidemiological parameters, and (iv) compare the rate of new cases arising from within-region transmission versus migration during the early epidemic.

## Materials and Methods

### Model

We fit a simplified version of the multi-type birth-death model described in (19). Under this model, beginning with a single infected host in a single geographic region (deme), the virus can be transmitted from one host to another (a birth event), die out due to host recovery or death (a death event), be sequenced (a sampling event, assumed to correspond to a death event), or migrate from one deme to another (a migration event). The birth, death, sampling, and migration processes are assumed to occur at deme-specific rates that are constant through time. Importantly, this model aims to capture heterogeneity in epidemiological parameters (birth and death rates) and sequencing effort (sampling proportion) among demes. We used a version of the model parameterized in terms of the effective reproductive number, which allows us to additionally infer this epidemiologically relevant quantity for each deme.

### Dataset

We analyzed SARS-CoV-2 genome sequences from five different demes: Hubei province in China, France, Germany, Italy, and a composite deme of other European countries (“other European”). All sequences were accessed from GISAID (12). To represent the pandemic origin, we randomly chose 10 sequences from Hubei collected on or before the lockdown of Wuhan city on 23 Jan. 2020. To investigate the earliest outbreaks in Europe, we considered all available sequences collected in France, Germany, and Italy on or before the lockdown of the Lombardy region of Italy on 8 Mar. 2020. These countries had the first detected (France and Germany) and the largest (Italy) early outbreaks in Europe (4, 7). By limiting sampling to before regional lockdowns and border closures went into effect, we hope to (i) satisfy model assumptions that epidemiological and migration parameters are constant through time, and (ii) get a picture of the early, unimpeded spread of SARS-CoV-2 within Europe. To represent the pool of SARS-CoV-2 circulating in other European countries during this time, we down-sampled sequences from other countries to the cumulative number of confirmed COVID-19 deaths in each country by 8 Mar. 2020 plus one (Table S1). We used this quantity as a proxy value roughly proportional to the outbreak size in each country. Table 1 characterizes the sequences analyzed from each deme for the main analysis. As a sensitivity analysis, we repeated the analysis while down-sampling based on confirmed death data from 28 Mar. 2020, considering that deaths occur with a delay after transmission. This yielded a slightly larger sequence set for analysis (results in supplement).

**Table 1.** Analyzed sequence information. Location is the location of sample collection, as recorded in the Nextstrain metadata (13). Date is the date of sample collection, as given on GISAID (12). No. = number; Seq. = sequence.

| Deme | No. seqs. | Locations represented | First seq. date | Last seq. date |
| --- | --- | --- | --- | --- |
| Hubei | 10 | Hubei province, China | 26.12.2019 | 18.01.2020 |
| France | 66 | France | 23.01.2020 | 08.03.2020 |
| Germany | 15 | Germany | 28.01.2020 | 03.03.2020 |
| Italy | 13 | Italy | 29.01.2020 | 04.03.2020 |
| Other European | 41 | Spain (18), Netherlands (4), United Kingdom (4), Switzerland (3), Belgium (1), Czech Republic (1), Denmark (1), Finland (1), Iceland (1), Ireland (1), Luxembourg (1), Norway (1), Poland (1), Portugal (1), Slovakia (1), Sweden (1) | 07.02.2020 | 08.03.2020 |

### Alignment generation

We prepared a sequence alignment from publicly available data on GISAID (12) using the Nextstrain pipeline for SARS-CoV-2 (13). Short sequences (< 25,000 bases), those without fully specified collection dates, and duplicate sequences from the same case were eliminated, as well as sequences from known transmission clusters or with suspicious amounts of nucleotide divergence (as determined by the Nextstrain team). We aligned selected sequences to reference genome Genbank accession MN908947. To eliminate suspected sequencing errors, we masked the first 130 and final 50 sites from the alignment, as well as sites 18,529, 29,849, 29,851, and 29,853 (following the Nextstrain pipeline).

### Testing assumptions about source and sink locations

We assume that during the time span considered here, the outbreak in Hubei and the different European outbreaks were only sources and not sinks for SARS-CoV-2 globally. In other words, we assume that (i) once a strain was in Europe, the strain could have been transmitted from Europe to other global regions, but subsequent re-introductions of this strain did not occur. Similarly, we assume (ii) strains were not re-introduced into Hubei. These assumptions allow us to ignore sequences from outside of Hubei and Europe. To justify assumption (ii), we argue there was not sufficient time between the pandemic origin in Hubei and Jan. 23, 2020 for a significant amount SARS-CoV-2 export, transmission outside-Hubei, and subsequent re-introduction into Hubei. Furthermore, confirmed case data shows that Hubei province was the epicenter of the SARS-CoV-2 pandemic until this time, with comparatively less transmission occurring outside of the province than within it (4). To justify assumption (i), we tested whether there was evidence for significant migration into European demes by running a separate analysis on A2a SARS-CoV-2 sampled from all global regions (results in supplement).

### Parameter inference

For inferences, we used the implementation of the multi-type birth-death model in the *bdmm* package (19, 20) in the BEAST2 software (21). Since this is a parameter-rich model, we fixed some parameters to improve the identifiability of others. The values for fixed parameters, priors for estimated parameters, and the rationale behind these decisions are given in Table 2. We ran four MCMC chains to approximate the posterior distribution of the model parameters. The first 10% of samples from each chain were discarded as burn-in before samples from the chains were pooled. We used Tracer (22) to assess the convergence and confirm that ESS was > 200 for all parameters.

**Table 2.** Values and priors for the parameters of the multi-type birth-death model. Confirmed case data for Hubei came from Statistica (24), for Germany, France, and Italy from the World Health Organization (4), and for other European countries from the European Center for Disease Control (25). The number of analyzed sequences divided by the number of confirmed cases provides an upper bound to the sampling proportion since confirmed cases are only a fraction of total cases. No. = number; approx. = approximately; IQR = inter-quartile range.

| Parameter | Value or Prior | Rationale |
| --- | --- | --- |
| Nucleotide substitution model | HKY + $\Gamma$ | Unequal transition/transversion rates, unequal base frequencies, rate heterogeneity among sites |
| Clock rate | 0.0008 | Approx. 24 mutations/year (13) |
| Death rate | 36.5 year<sup>-1</sup> | Period between infection and becoming un-infectious assumed exponentially distributed with a mean of 10 days |
| Sampling start time | 23 Dec. 2019 | Just before date of first sample |
| Sampling end time (Hubei only) | 23 Jan. 2019 | Only included sequences collected until lockdown |
| Location of origin | Hubei | Putative pandemic origin |
| Reproductive number | Lognormal(0.8, 0.5) | Median 2.3, 95% IQR 0.8 – 5.9 |
| Migration rates | Lognormal(0, 1) | Median time until travel is 1 year, 95% IQR 51 days – 7.1 years |
| Time of origin | Lognormal(-1, 0.2) | Median 26 Oct., 95% IQR 22 Aug. – 8 Dec. 2019 |
| Sampling proportion |  | Upper bounds based on confirmed cases: |
| Hubei | Uniform(0, 0.15) | 10/66 cases on 18 Jan. 2020 |
| France | Uniform(0, 0.093) | 66/706 cases on 8 Mar. 2020 |
| Germany | Uniform(0, 0.10) | 15/157 cases on 3 Mar. 2020 |
| Italy | Uniform(0, 0.005) | 13/2,502 cases on 4 Mar. 2020 |
| other European | Uniform(0, 0.057) | 41/712 cases on 8 Mar. 2020 |

### Comparing rates of migration and within-region transmission

To weigh the significance of cases from migration versus within-region transmission during the early epidemic, we compare the rate at which new cases migrate into a region (= per-individual migration rate x case count in source region) to the rate at which new cases arise from within-region transmission (= transmission rate x case count in sink region). When signal in the sequence data is low, e.g. for some migration rates, our prior assumptions determine the magnitude of these rates. To assess the sensitivity of our main conclusions to the prior, we additionally analyzed the same sequences using a lower migration rate prior (Figure S12B). We note that the migration and transmission rates are assumed to be constant through time for this analysis. Thus, the temporal trends depend only on the confirmed case data, which we take from the Johns Hopkins Center for Systems Science and Engineering (23).

## Results

### Testing assumptions about source and sink locations

To test our assumption that Europe was primarily a source and not a sink of infections before 8 Mar. 2020, we analyzed A2a sequences collected from different global regions on or before that date. We aggregated sequences into five demes: Africa, Asia & Oceania, Europe, North America, and South & Central America (Table S3), and then fit the multi-type birth-death model to these data. The most recent common ancestor of the global set of A2a sequences was inferred to be in Europe with 95% posterior support (Figure S9). The posterior distributions for the migration rates into Europe closely matched the prior, thus the data contains little information on these rates (Figure S10). However, in the analyzed dataset, 0 introduction events were inferred from other parts of the world into Europe, while in total 24 migration events were inferred from Europe to other parts of the world (Table S5).

### Inference results: SARS-CoV-2 transmission into and across Europe

For our main analysis we focused on estimating patterns of SARS-CoV-2 transmission into and across Europe. Based on the particular set of sequences analyzed, we infer that SARS-CoV-2 was introduced from Hubei into France, Germany, Italy and other European countries approximately 2-4 times each before 8 Mar. 2020 (Table 3). The largest number of estimated introductions was 18 from Italy to other European countries. Importantly, these estimates reflect only introductions occurring in the transmission history of the analyzed cases, not the full epidemic. In contrast, the inferred migration rate parameters should describe more general patterns of spread between regions. The sequence data were informative for inferring some, but not all, migration rates. We highlight here only the rates for which the data is the most informative; see Figure S1 for a full comparison of posterior and prior distributions. The highest migration rate was inferred to be from Italy into other European countries, with a median rate of 3.7/year. The lowest migration rate was from Italy to Germany, with a median rate of 0.44/year.

**Table 3.** Median inferred number of introductions from each source deme to each sink deme along the transmission tree linking analyzed cases. Hubei is assumed to be a source only. Values in brackets are the upper and lower bound of the 95% highest posterior density interval for these estimates.

| Source / sink | France | Germany | Italy | other European |
| --- | --- | --- | --- | --- |
| Hubei | 3: [1, 6] | 4: [1, 6] | 2: [0, 6] | 4: [0, 8] |
| France | - | 0: [0, 1] | 0: [0, 3] | 2: [0, 4] |
| Germany | 0: [0, 2] | - | 1: [0, 3] | 1: [0, 4] |
| Italy | 6: [1, 9] | 1: [0, 4] | - | 18: [7, 34] |
| other European | 2: [0, 6] | 1: [0, 4] | 1: [0, 4] | - |

### Inference results: A2a lineage origin

The maximum clade credibility tree in Figure 1 summarizes the posterior sample of transmission trees linking analyzed sequences. The A2a lineage sequences form a clear clade with posterior support of 1. The most recent common ancestor of the analyzed A2a sequences is estimated to be in Italy with 87% posterior support. In contrast, the location of the most recent common ancestor between this clade and the basal, Singapore-linked German sequence is less certain. This ancestor is inferred to have been in either Germany (40% posterior support), Hubei (38%), or Italy (20%). We find very little support for this ancestor having been in France or another European country (2%).

**Figure 1.**
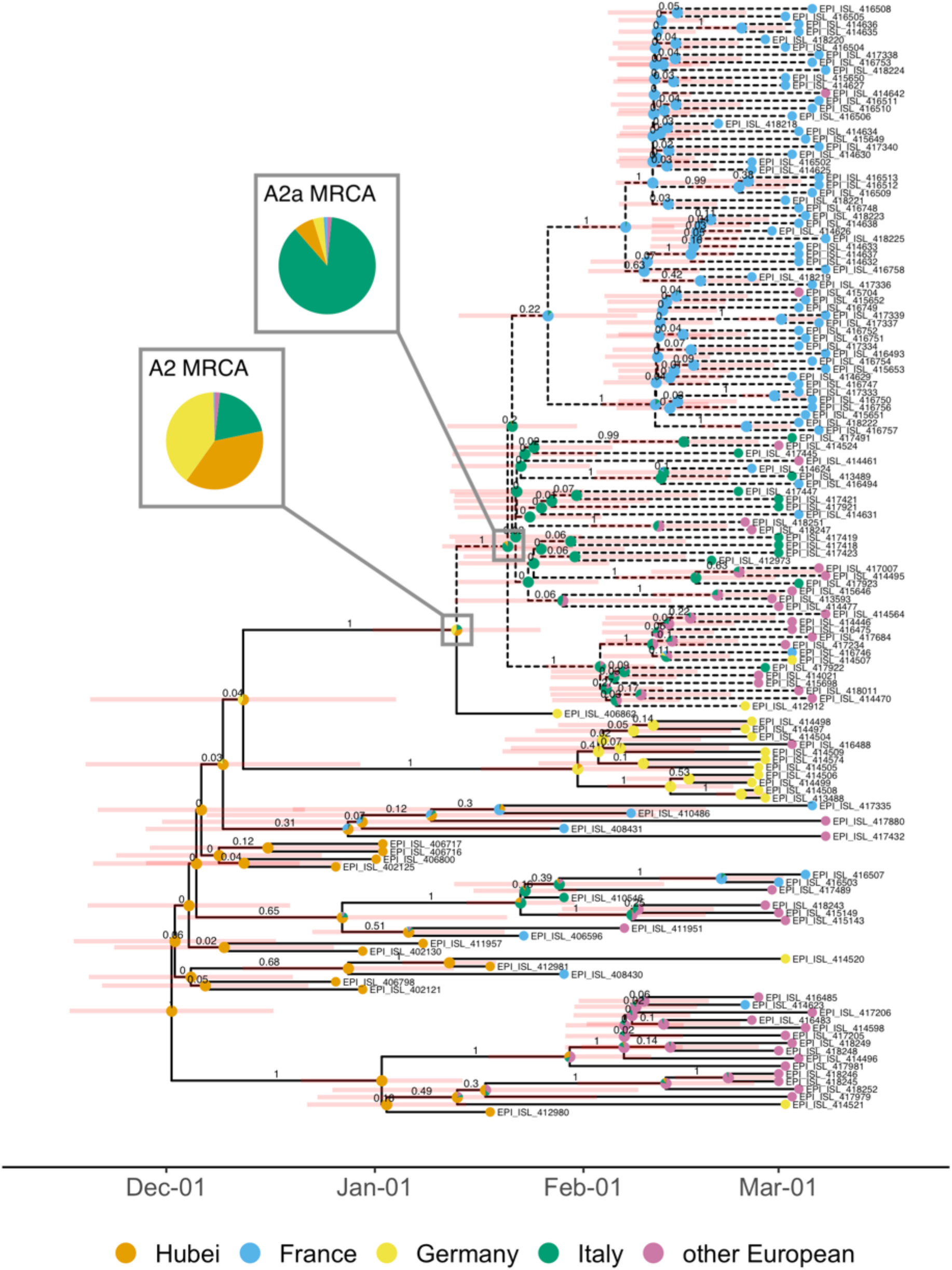
Maximum clade credibility tree. The clade of A2a sequences analyzed is highlighted with dashed branches. The values above the branches are the posterior clade probabilities and the pale red bars show the 95% highest posterior density interval for node ages. The pie charts at nodes show posterior support for the ancestor being located in each deme (note that we assumed the root of the tree was in Hubei with probability 1). The deme for each tip is the deme in which the sequence was collected, irrespective of travel history. Tips are annotated with GISAID accession identifier.

### Inference results: epidemiological parameters

Several epidemiologically relevant parameters were co-inferred along with the transmission tree. Firstly, we report on the reproductive number in the different demes, which varied from 1.2 to 1.7 in Hubei to 2.5 to 3.5 in France (Figure S2). Secondly, we report on the prevalence of no-longer infectious cases in each deme as of the collection date of the last analyzed sequence. This quantity can be back-calculated from the estimated sampling proportion (prevalence = # sequences analyzed / sampling proportion). We note that both the sampling proportion and prevalence estimates have large credible intervals (Figures S3 and S4). Of the European demes analyzed, the outbreak in Germany was estimated to be smaller in early March (150 to 490 cumulative cases) than the outbreaks in France (709 to 2,323 cases) and other European countries (719 to 1,806 cases), while the outbreak in Italy was the largest (2,600 to 4,988 cases).

### Comparing rates of migration and within-region transmission

Figure 2 compares the rate at which we estimate new cases to arise in each region from migration versus from within-region transmission. The estimated rates of new cases from migration and within-region transmission are represented here as point estimates 5 days before the date of case confirmation, which assumes a 5-day delay between infection and onward transmission or migration. Beginning with the first day on which we have case data from Hubei, we estimate a substantial risk of infected individuals migrating from Hubei into European regions. Throughout late January to mid-February 2020, cases were sporadically detected in each European region, each of which is associated with a risk of subsequent within-region transmission. Sustained within-region transmission is first evident in Italy in mid-February. Shortly thereafter, sustained within-region transmission occurred in other European countries, in France, and in Germany. By 8 Mar. 2020, the estimated rate of occurrence of new cases from within-region transmission is within the estimated bounds on the rate of new cases from migration for each region considered (Figure S12A). We obtain the same qualitative result in our sensitivity analysis using a very different prior on the migration rate (Figure S12B). We note that the rates in Figure 2 are underestimates of the rates of new cases arising due to migration or transmission due to the underreporting in the confirmed case data. However, assuming that the amount of underreporting is comparable across regions, we can indeed compare the rates.

**Figure 2.**
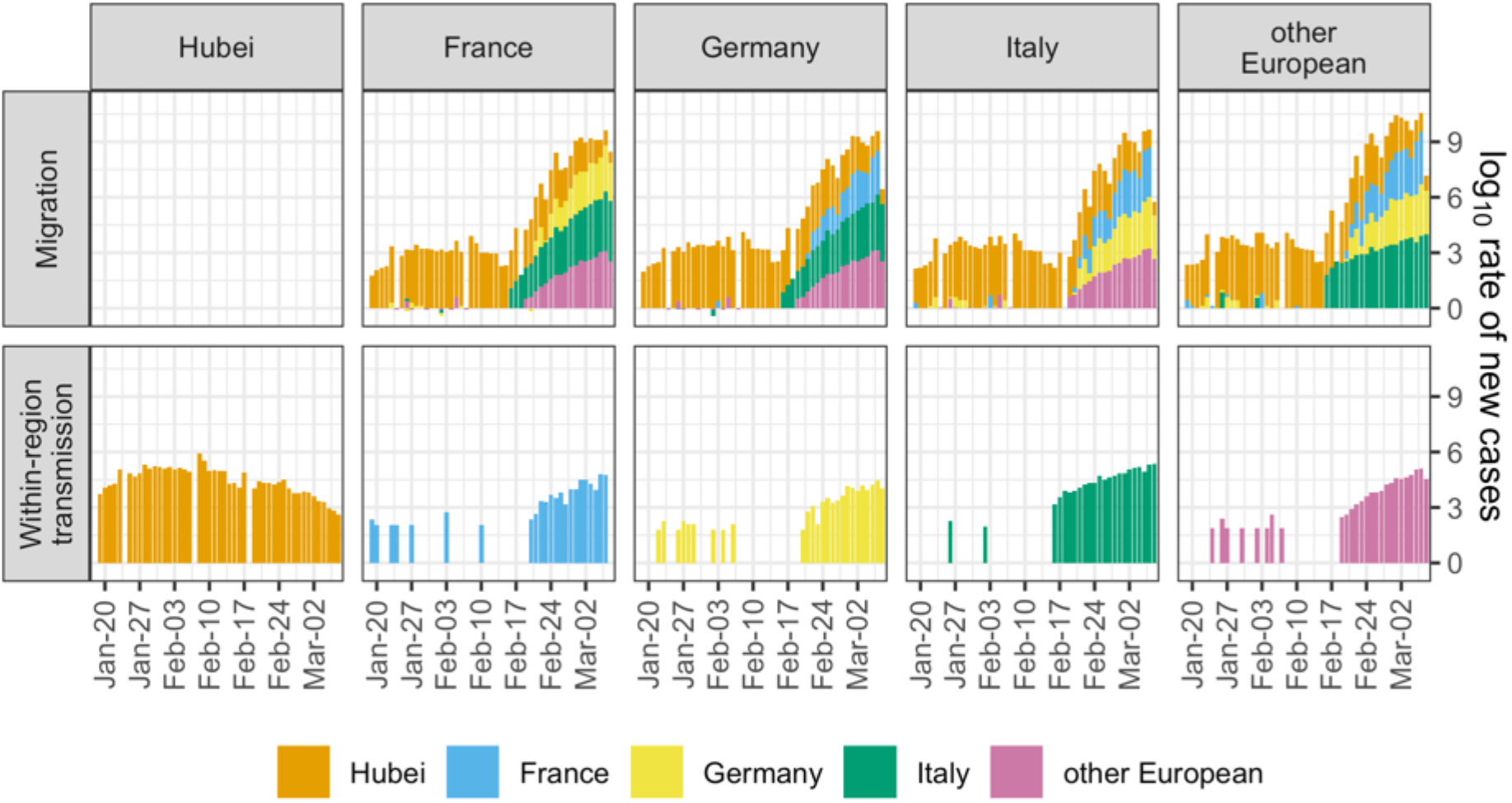
Estimated rate of new cases arising from migration compared with the estimated rate of new cases arising from within-region transmission. For each day, we multiplied the number of newly confirmed cases in each source region by the posterior sample of migration rates from source to sink. The median of these rates is show in the “Migration” row. We also multiplied the number of newly confirmed cases in each sink region by the posterior sample of transmission rates for the region. The median of these rates is shown in the “Within-region transmission” row. Dates are lagged 5 days to account for a 5-day delay between infection and migration or onward transmission. Case data comes from (23).

## Discussion

We inferred the early spread of the SARS-CoV-2 virus into and across Europe as well as the geographic origin of the predominant A2a lineage spreading in Europe. To do this, we applied a previously published phylodynamic model to analyze publicly available viral genome sequences from the epidemic origin in Hubei, China and from the earliest detected and largest European outbreaks before 8 Mar. 2020. After performing Bayesian inference, we (i) report on inferred patterns of SARS-CoV-2 spread into and across Europe, (ii) compare posterior support for several hypotheses on the origin of the A2a lineage, (iii) report on epidemiological parameters, and (iv) compare the timeline of new cases resulting from migration versus within-region transmission in Europe before borders were closed.

Genome sequence data indicates that prior to 8 Mar. 2020, SARS-CoV-2 was introduced from Hubei province into France, Germany, Italy and other European countries at least 2-4 times each (Table 1). These estimates, which are based solely on genome sequence data, provide a complementary account of introduction events compared to line-list data (26). The introduction events we report here are inferred to have occurred along the transmission tree specific to the analyzed sequence set and are not attributable to individual cases. In comparison, line-list data (7, 26) attributes introduction events to individual cases but cannot reconstruct previous, unobserved introductions. Since we analyze only a fraction of all cases, we expect our estimates to be a minimum bound on the true number of introductions.

Ideally, we want to go beyond counting migration events amongst the analyzed sequences and investigate general dynamics. To do this, we would interpret inferred migration rates as representing more general patterns of SARS-CoV-2 spread. However, the sequence data was only informative for inferring some of these rates (Figure S1). In regions with few lineages circulating during the period considered, there is little signal for the amount of outward migration. We observe information about the per-individual migration rate from Italy to other European countries (Figure S1). However, we do not find evidence for preferential migration paths from Hubei into different European regions or from each European region to the others, although we cannot exclude this possibility.

We estimate that the A2a viruses spreading in Europe by 8 Mar. 2020 had a common ancestor in Italy sometime between mid-January and mid-February 2020 (Figure 1). In contrast, Nextstrain places this ancestor in the U.K. with 100% confidence (27). However, the Nextstrain result may be an artefact of disproportionately high sequencing effort in the U.K. since biased sampling violates the assumptions of the “mugration” method employed (28). We additionally report that the A2a lineage was most likely carried from Hubei to Italy or from Hubei to Italy via Germany. Both transmission routes have almost equal posterior support under our model assumptions (Figure 1). Since we only consider a few geographic regions, these migration routes are not necessarily comprehensive. Rather than reconstructing a complete transmission chain, we compare model support for different A2a transmission routes amongst the analyzed demes and report two equally plausible routes.

Although it is not the main focus of our analysis, we also report on epidemiological parameters of the early outbreaks considered. Estimates for the reproductive number fall roughly within the range of previous estimates (29), though we mention a particular caveat with respect to the reproductive number in Hubei below. Unsurprisingly, prevalence estimates in early March generally exceed confirmed case counts by a factor of 1-3 (Figure S4). Our inferences of epidemiological parameters do not challenge the idea that the early reproductive number in different outbreaks is difficult to estimate precisely, but not hugely variable, and that there is substantial under-reporting in line-list data (30).

Finally, we estimated the rate of new cases arising from migration compared with the rate of new cases arising from within-region transmission in the regions analyzed. The magnitudes of these rates are quite uncertain due to uncertainty in the inferred migration and transmission rates (Figure S11) and under-reporting in case counts, which we implicitly assume to be constant in time and between demes. However, the temporal trends suggested by these data are still compelling and robust towards different prior assumptions. We see that under sustained risk of case migration from Hubei, isolated cases were confirmed throughout Europe beginning in late January 2020 but did not immediately cause large outbreaks. Shortly after the first evidence of sustained within-region transmission in Italy, outbreaks in the rest of Europe also took hold (Figure 2).

Our results based on the multi-type birth-death model take into account phylogenetic uncertainty and sampling biases between demes, which are two major concerns in genomic analyses of SARS-CoV-2 (18). Indeed, wide confidence intervals around internal nodes in the maximum clade credibility tree and low clade support near the tips (Figure 1) indicate a high degree of phylogenetic uncertainty. Therefore, it is important that the parameter estimates we report result from integrating over a distribution of potential phylogenies with different geographic locations assigned to ancestral lineages. In comparison, some initial studies that estimated international SARS-CoV-2 spread constructed a median-joining network instead of a phylogeny to account for this uncertainty (16, 31). In this approach, identical sequences are collapsed to single nodes and edges represent mutational differences. This disregards information from relative sampling times and means that ancestor-descendent relationships are highly dependent on the choice of the network root (32, 33). Unaccounted-for sampling biases in these analyses may also yield spurious results for the geographic origin of lineages (34, 35). Our analysis, which relies on a mechanistic model of migration and between-deme sampling differences, should be robust to such biases.

Despite the advantages of the multi-type birth-death model just mentioned, there are also several unique caveats to consider. The birth-death model assumes uniform-at-random sampling from the total infected population in each deme. However, particularly in the early stages of outbreaks, infected individuals were identified by health ministries via contact tracing (7). Non-random sampling may be one possible explanation for why we infer markedly different transmission rates in China when analyzing cases from within Hubei (as in this analysis) as opposed to cases exposed in Hubei but sequenced elsewhere (as in our previous analysis (36)). Furthermore, the multi-type birth-death model assumes that parameters are constant through time and homogenous within demes. As a result, our inferences based on province-, country-, and continent-level demes are only coarse approximations of the true, heterogeneous epidemic dynamics occurring at a local level. In particular, we do not account for a reduction in airline traffic between Europe and China beginning in late January, before borders were closed (37). Due to these limitations, we focus on estimating and interpreting particular events along the transmission tree of the analyzed sequences (e.g. Table 3, Figure 1) and advise caution when interpreting inferred migration rates (Figure S1).

We expect that our results will be useful in parameterizing more specialized models aimed to understand the efficacy of border closures as a means to fight pandemic disease. So far, such analyses have primarily used line-list data and information on travel networks to estimate SARS-CoV-2 migration patterns (38–40). Here we present independent estimates of migration patterns based on genome sequence data. By combining case count data and our estimates for migration and transmission rates, we provide a timeline of early SARS-CoV-2 introduction and spread before border closures were implemented. Despite migration risk from Hubei being on the same order of magnitude as later migration risk from Italy, we only observe sustained outbreaks in other European regions after the onset of sustained within-region transmission in Italy. Finally, before the first border closures in Europe, we estimate the risk of new cases arising from within-region transmission to be within the estimated range for the risk of new migration cases.

## Data Availability

The data used for this manuscript is publicly accessible on GISAID.org. We have included an acknowledgments table detailing which data were used for each analysis.

https://www.gisaid.org/

## Acknowledgments

S.N, T.V., J.S., J.H., and T.S. thank ETH Zürich for funding.

